# Effect of Renin-Angiotensin System (RAS) Inhibitors in Acute Ischemic Stroke to Improve Outcomes in In-Patient Settings: A Cross Sectional and Longitudinal Analysis

**DOI:** 10.1101/2023.12.05.23299561

**Authors:** Sophie Samuel, Kyndol Craver, Charles Miller, Brittany Pelsue, Catherine Gonzalez, Teresa A Allison, Brian Gulbis, H Alex Choi, Seokhun Kim

## Abstract

**Background:** Acute ischemic stroke (AIS) is a major health challenge, often resulting in long-term disability and death. This study assesses the impact of renin-angiotensin system (RAS) inhibitors (angiotensin-converting enzyme inhibitors and angiotensin receptor blockers) on AIS patient mortality compared to non-RAS antihypertensive medications.

**Methods:** This retrospective cohort study, conducted at Memorial Hermann–Texas Medical Center in Houston, Texas, from August 31, 2017, to August 31, 2022, examined AIS patient mortality. We used a cohort design, evaluating the effects of RAS inhibitors, either alone or in combination with beta-blockers (BBs), while exploring interactions, including those related to end-stage renal disease (ESRD) and serum creatinine levels. Eligible subjects included AIS patients aged 18 or older with specific AIS subtypes who received in-patient antihypertensive treatment. Missing data were addressed using imputation, followed by Inverse Probability of Treatment Weighting (IPTW) to achieve covariate balance. Our primary outcome was mortality rates. Statistical analyses involved cross-sectional and longitudinal approaches, including generalized linear models, G-computation, and discrete time survival analysis over a 20-day follow-up period.

**Results:** In our study of 3058 AIS patients, those using RAS inhibitors had significantly lower in-hospital mortality (2.2%) compared to non-users (12.1%), resulting in a relative risk (RR) of 0.18 (95% CI 0.12-0.26). Further analysis using G-computation revealed a marked reduction in mortality risk associated with RAS inhibitors (Risk 0.0281 vs. 0.0913, Risk Difference (RD) of 6.31% or 0.0631, 95% CI 0.046-0.079). Subgroup analysis demonstrated notable benefits, with individuals having creatinine levels below and above 1.3 mg/dL exhibiting statistically significant RD (RD −0.0510 vs. −0.0895), and a significant difference in paired comparison (−0.0385 or 3.85%, CI 0.023-0.054). Additionally, longitudinal analysis confirmed a consistent daily reduction of 0.93% in mortality risk associated with the intake of RAS inhibitors.

**Conclusion:** RAS inhibitors are associated with a significant reduction in in-hospital mortality in AIS patients, suggesting potential clinical benefits in improving patient outcomes.

## Introduction

Acute ischemic stroke (AIS), a leading cause of long-term functional disability and mortality in adults, often presents with elevated blood pressure, which, if uncontrolled, can lead to poorer outcomes.^1–3^ Antihypertensives are a key therapeutic option in the ischemic stroke regimen to manage blood pressure and improve outcomes.^3–7^

Given the significance of blood pressure control in AIS, the potential benefits of renin-angiotensin system (RAS) inhibitors have gained attention.^7–11^ Several pivotal trials, including the HOPE trial, which demonstrated angiotensin (ACE) inhibition’s positive effects on cardiovascular events and mortality, have contributed to this exploration.^12^ The ONTARGET and TRANSCEND trials have similarly highlighted the cardiovascular benefits of angiotensin-receptor blockers (ARBs) in high-risk patient populations.^7,12,13^ The utilization of RAS inhibitors in stroke patients has been studied in trials. The ACCESS trial evaluated a 7-day course of candesartan post-acute ischemic stroke, which significantly improved cardiovascular morbidity and mortality. ^14^

In patients with end-stage renal disease (ESRD), RAS inhibitors have also been shown to have benefits in reducing cardiovascular events and mortality.^15–17^ A meta-analysis of randomized controlled trials found that ACE inhibitors and ARBs reduced all-cause mortality, cardiovascular mortality, and major cardiovascular events in patients with ESRD.^18^ However, the use of RAS inhibitors in patients with high creatinine levels or acute kidney injury (AKI) has been a topic of debate. While some studies suggest that RAS inhibitors may exacerbate AKI, other studies have shown no increased risk of AKI or mortality in patients with high creatinine levels. ^19–21^

Mechanistically, RAS inhibitors have been shown to have neuroprotective effects and improve cerebral blood flow, while beta blockers (BBs) can reduce sympathetic nervous system activity and improve cardiac function.^22,23^ A systematic review and meta-analysis by Strauss et al. showed that RAS inhibitors and BBs have a synergistic effect in reducing cardiovascular and renal outcomes in patients with type 2 diabetes.^24^ Furthermore, a meta-analysis by Wiysonge et al. demonstrated the neuroprotective effects of BBs in AIS patients.^25^ Combining RAS inhibitors and BBs may lead to improved blood pressure control, reduced inflammation, and improved cerebral perfusion. These studies support the hypothesis that combining RAS inhibitors and BBs could lead to improved outcomes in AIS patients.

Our study aims to assess the impact of RAS inhibitors on mortality in AIS patients, investigating interactions with BBs, ESRD, and creatinine levels. We aim to determine whether RAS inhibitors (ACE inhibitors or ARBs) reduce mortality compared to other antihypertensive medications in AIS patients, examining both cross-sectional and longitudinal effects.

## Methods

### Study design and settings

This retrospective observational study was conducted at Memorial Hermann–Texas Medical Center, a large academic medical center in Houston, Texas.

### Patient population

Our study included all adults admitted to Memorial Hermann Hospital – Texas Medical Center with AIS between August 31, 2017, and August 31, 2022. Inclusion criteria were adults with AIS symptoms, confirmed by head CT imaging, a GCS score of 5 or higher at ED arrival, and a Trail of Org 10172 in Acute Stroke Treatment (TOAST) classification of larger vessel occlusion, small vessel disease, or cardioembolic stroke, who received in-patient acute blood pressure control with antihypertensives. Exclusions applied to cases with uncommon stroke etiologies or missing data in the University of Texas REGIS database.

### Intervention and comparison

Patients received either RAS inhibitors (ACE inhibitors or ARBs) or non-RAS antihypertensives, including BBs, CCBs, and any other antihypertensive medications. This observational study considers daily variation in RAS inhibitor intake, necessitating a longitudinal analysis to assess their impact on mortality. Data was censored at day 20 to match the study timeframe. For cross-sectional analysis, patients were categorized into two groups: ‘RAS’ (RAS medication for at least one day) vs. ‘non-RAS’ (no RAS medication). We also examined interactions between RAS inhibitors, BBs, ESRD, creatinine levels, and their impact on mortality.

### Outcomes

The primary outcome of this study was mortality, aiming to assess the impact of RAS inhibitors on mortality in AIS patients. We specifically examined the interaction between RAS inhibitors and subgroups categorized by BBs, ESRD, and creatinine levels, considering both daily mortality for longitudinal analysis and aggregated mortality for cross-sectional analysis.

### Covariates

In our analysis, we considered a comprehensive set of baseline covariates, including beta blockers, serum creatinine, age, NIHSS score, history of ESRD, the presence of large vessel occlusion, small vessel disease, weight, pre-morbid mRS, history of ischemic stroke, history of type 2 diabetes, history of hypertension, baseline systolic pressure, history of atrial fibrillation, ethnicity, and race. Additionally, we included patients’ home RAS medication, time-varying daily RAS intake (measured at the previous day), and time-varying daily creatinine levels as covariates for the longitudinal analysis.

### Statistical analysis

At the outset of our study, we initiated the research by conducting comprehensive descriptive and preliminary bivariate analyses. We employed MissForest,^26,27^ a machine learning method known for its accuracy and efficiency in imputing mixed-type (continuous and categorical) missing data with complex relations. This was particularly relevant for our study, which had a large master database with missing data for approximately 5-10% of the data points. The distribution of missing values across the baseline variables varied. Past medical history variable exhibited the maximum missing percentage, close to 10%, while other baseline variables, including baseline blood pressure readings, showed relatively lower percentages of missing data, ranging from 5% to 8%. Additionally, less than 10% of the data was missing in daily creatinine levels within the longitudinal dataset, whereas baseline creatinine levels were complete. Notably, all mortality values and RAS intake values were reported on a daily basis without any missing data.

### Inverse probability weighting (IPW)

We utilized inverse probability of treatment weighting (IPTW) to adjust for confounding by imbalanced covariates.^28^ For cross-sectional analysis, we employed a logistic regression model to calculate the propensity score for cross-sectional RAS inhibitor intake, derived from the baseline covariates. We assessed the performance of the model through diagnostic plots and balance statistics, with a predefined threshold of < 0.25 in standardized mean differences. For longitudinal analysis, we used Gradient boosting machines (GBMs)^29,30^ to estimate the propensity score of time-varying RAS inhibitor intake at each day from the baseline covariates, prior use of RAS as home medication, RAS inhibitor intake at the previous day, and creatinine level at the same day, to deal with relatively higher correlation among those time-varying variables. Moreover, in order to handle potential informative censoring, inverse probability of censoring weights (IPCW) were calculated by estimating the probability of remaining up to each day using GBMs with the same covariates (except that creatinine level at the previous day was used instead of at the same day). IPTW and IPCW were multiplied to obtain the final weights for each day.

### Cross-sectional analysis

A logistic regression with IPTW was used to model the relationship between RAS and mortality. All the baseline covariates were included in the model to achieve a doubly-robust estimate of the RAS effect. ^30^ Further, we also examined potential interactions between RAS and other predictors in the model. Specifically, we included interaction terms between RAS and BBs, baseline serum creatinine, and history of ESRD to investigate potential subgroups who may benefit differently from RAS.

Due to non-collapsibility of logistic regression, however, the coefficient of RAS effect in the model is not the same as its marginal effect, which is an unconditional average treatment effect (ATE) of RAS. Thus, G-computation^31,32^ method was adopted, by calculating weighted average potential outcomes for each counterfactual scenarios (i.e., all receiving RAS vs. all not receiving RAS) and obtaining their contrast (e.g., risk difference) as the estimate of treatment effect, which are unconditional on covariates in the model. Further, G-computation was applied targeting each effect modifier separately, to obtain an ATE in each subgroup of the effect modifier.

### Longitudinal analysis

After completing the initial cross-sectional analysis, we proceeded with a rigorous longitudinal data analysis to delve deeper into temporal dynamics. Given that the longitudinal data is based on discrete time intervals (i.e., days), we employed a multiperiod logit model^33–35^, which is a discrete time survival model, with the final weights (i.e., IPTW x IPCW) being applied, to obtain an unbiased estimate of time-dependent RAS effect on mortality. The model also included daily serum creatinine levels (at the same day), daily use of RAS medications (the previous day), and the presence of home RAS medications as covariates to adjust for their remaining imbalances and achieve double-robustness. Finally, a G-computation method was applied for the fitted model to obtain an unconditional ATE of daily RAS intake. We performed all statistical analysis using R version 4.0.2 (R Foundation for Statistical Computing, Vienna, Austria) and utilized the “WeightIt” package for IPW, the “marginal effects” package for G-computation, and “dynamichazard” package for discrete time survival modeling. We considered p-values less than 0.05 to be statistically significant using a two-tailed test. We employed various statistical methods to account for potential confounding in our observational study.

## Results

### Preliminary Data Analysis

The baseline characteristics of the patients in the RAS inhibitor group (n = 1576) and the non-RAS inhibitor group (n = 1482) were imbalanced prior to the application of missing imputation and IPTW. Table 1 shows that patients in the RAS inhibitor group were more likely to be Black or African American and Hispanic or Latino, while patients in the non-RAS inhibitor group were more likely to be White/Caucasian and non-Hispanic. Additionally, patients in the RAS inhibitor group had a slightly lower mean NIHSS score (8.63 vs 10.3) and were more likely to have a pre-morbid mRS score of 0-2 (70.1% vs 65.3%). Moreover, patients in the RAS inhibitor group were more likely to have small vessel occlusion (18.8% vs 10.7%). Notably, the RAS inhibitor group had a slightly lower median baseline serum creatinine (1.05 vs 1.07) and a lower proportion of patients with

**Table 1.** Baseline Characteristics prior to missing imputation and IPTW. Afib = atrial fibrillation, BB = beta blocker, BP = blood pressure, CAD = coronary artery disease, CCB = calcium channel blocker, CHF = congestive heart failure, DVT = deep venous thromboembolism, ESRD = end stage renal disease, EVT = Endovascular therapy, ICH = intracerebral hemorrhage, MI = myocardial infarction, Misc = miscellaneous, PE = pulmonary embolism, tPA = tissue plasminogen activator, T1DM = type 1 diabetes mellitus, T2DM = type 2 diabetes mellitus, UTI = urinary tract infection

| Characteristic | RAS inhibitors<br>n = 1576 | non-RAS<br>inhibitors<br>n= 1482 | p-value |
| --- | --- | --- | --- |
| **Age (years) - mean (SD)** | 66.4 (13.1) | 67.2 (15.0) | 0.094 |
| **Female, n (%)** | 679 (43.1) | 672 (45.3) | 0.261 |
| **Weight (kg) – mean (SD)** | 84.9 (23.9) | 82.8 (22.6) | 0.014 |
| **Race, n (%)** |  |  |  |
| White/Caucasian | 509 (32.3) | 569 (38.4) | <0.001 |
| Black or African American | 489 (31.0) | 396 (26.7) | 0.009 |
| Asian | 37 (2.3) | 39 (2.6) | 0.614 |
| **Ethnicity, n (%)** |  |  |  |
| Hispanic or Latino | 288 (18.3) | 231 (15.6) | 0.048 |
| Non-Hispanic | 1228 (77.9) | 1200 (81.0) | 0.037 |
| **National Institutes of Health Stroke Scale (NIHSS ) - mean (SD)** |  |  |  |
| NIHSS score – mean (SD) | 8.63 (7.49) | 10.3 (9.03) | <0.001 |
| **National Institutes of Health Stroke Scale (NIHSS) – n (%)** |  |  |  |
| NIHSS Score 0 - n (%) | 124 (7.9) | 115 (7.8) | 0.911 |
| NIHSS score 1 - 4 - n (%) | 457 (29.0) | 389 (26.2) | 0.089 |
| NIHSS score 5 - 15 - n (%) | 626 (39.7) | 475 (32.1) | <0.001 |
| NIHSS score 16 - 20 - n (%) | 151 (9.6) | 178 (12.0) | 0.030 |
| NIHSS score 21 - 42 - n (%) | 137 (8.7) | 236 (15.9) | <0.001 |
| **Pre-Morbid Modified Rankin Scale (mRS)** |  |  |  |
| mRS score 0-2 n(%) | 1107 (70.1) | 968 (65.3) | 0.004 |
| mRS score 3-5 n(%) | 257 (16.3) | 293 (19.8) | 0.013 |
| **TOAST Acute Stroke Etiology - n (%)** |  |  |  |
| Large artery atherosclerosis | 440 (27.9) | 395 (26.7) | 0.432 |
| Small vessel occlusion | 296 (18.8) | 159 (10.7) | <0.001 |
| Cardioembolic | 841 (53.4) | 930 (62.8) | <0.001 |
| **Admission Blood Pressure- mean(SD)** |  |  |  |
| Systolic Blood Pressure, mmHg | 163 (32.3) | 149 (29.7) | <0.001 |
| Diastolic Blood Pressure, mmHg | 87.7 (18.5) | 82.1 (24.3) | <0.001 |
| **Initial presentation – n (%)** |  |  |  |
| In-Hospital | 140 (8.9) | 170 (11.5) | 0.018 |
| Direct presentation | 728 (46.2) | 617 (41.6) | 0.011 |
| Transfer from other hospital | 708 (44.9) | 695 (46.9) | 0.274 |
| **Baseline creatinine – median (IQR)** |  |  |  |
| Serum creatinine , mg/dL | 1.05 (0.84-1.32) | 1.07 (0.85-1.50) | <0.001 |
| **Dialysis – n (%)** |  |  |  |
| CRRT | 32 (2.0) | 75 (5.1) | <0.001 |
| IHD | 43 (2.7) | 96 (6.5) | <0.001 |
| Peritoneal | 2 (0.1) | 9 (0.6) | 0.027 |
| **Past Medical History - n (%)** |  |  |  |
| Ischemic Stroke | 267 (17.9) | 232 (15.7) | 0.272 |
| Atrial Fibrillation | 309 (20.3) | 343 (23.1) | 0.019 |
| ESRD | 38 (2.4) | 63 (4.3) | 0.005 |
| HTN | 1266 (84.2) | 1043 (70.4) | <0.001 |
| **Stroke Course - n (%)** |  |  |  |
| tPA Administered - n (%) | 374 (23.7) | 355 (24.0) | 0.893 |
| EVT performed - n (%) | 241 (18.4) | 236 (15.9) | 0.515 |
| **General complications During Hospitalization – n (%)** |  |  |  |
| Hemorrhagic transformation complication | 268 (17.0) | 194 (13.1) | 0.003 |
| **Medication administered breakdown- n (%)** |  |  |  |
| BB only | 295 (18.7) | 648 (43.7) | <0.001 |
| BB and CCB | 157 (10.0) | 207 (13.9) | <0.001 |
| BB and Misc BP meds | 115 (7.3) | 39 (2.6) | <0.001 |
| BB, CCB, and Misc BP meds | 265 (16.8) | 124 (8.4) | <0.001 |
| BB, Diuretics, and Misc BP meds | 65 (4.1) | 32 (2.2) | 0.002 |
| BB, CCB, diuretics and Misc BP meds | 233 (14.8) | 105 (7.1) | <0.001 |
| CCB only | 32 (2.0) | 37 (2.5) | 0.386 |
| CCB and diuretics | 8 (0.5) | 9 (0.6) | 0.711 |
| CCB and Misc BP meds | 71 (4.5) | 25 (1.7) | <0.001 |
| Diuretics only | 10 (0.6) | 8 (0.5) | 0.732 |
| Misc BP meds only | 24 (1.5) | 14 (0.9) | 0.149 |
| Diuretics and Misc BP Meds | 6 (0.4) | 5 (0.3) | 0.841 |
| RAS inhibitors only | 31 (2.0) | 0 (0.0) | <0.001 |
| No BP medication | 0 (0.0) | 37 (2.5) | <0.001 |
| **Other medications – n (%)** |  |  |  |
| Vasopressors | 388 (24.6) | 474 (32.0) | <0.001 |
| Nephrotoxic agents | 199 (12.6) | 217 (14.6) | 0.104 |
| **Clinical outcome – n (%)** |  |  |  |
| Mortality - n (%) | 34 (2.2) | 179 (12.1) | <0.001 |

ESRD (2.4% vs 4.3%). Furthermore, patients in the non-RAS inhibitor group had a slightly lower mean weight (82.8 kg vs 84.9 kg) and were more likely to receive Continuous Renal Replacement Therapy (CRRT) and Intermittent Hemodialysis (IHD). Moreover, patients in the non-RAS inhibitor group were more likely to receive BB only and BB with CCB, while patients in the RAS inhibitor group were more likely to receive a combination of BB, CCB, diuretics, and miscellaneous BP medications. (Table 1)

### Preliminary Data Outcomes

Significant differences were observed between patients treated with RAS inhibitors and those without, particularly in terms of all-cause in-patient mortality. The non-RAS inhibitor group exhibited a notably higher mortality rate (12% vs. 2%, p < 0.001).

### Inverse probability weighting and covariate balances

For cross-sectional analyses, IPTW successfully achieved balance of covariates between the RAS inhibitor and non-RAS inhibitor groups with standardized mean difference (SMD) of 0.25 as the criterion. The SMD column in figure 3a shows the degree of balance achieved between treatment and control groups on each covariate after propensity score balancing, with smaller values indicating better balance.

**Figure 3a.**
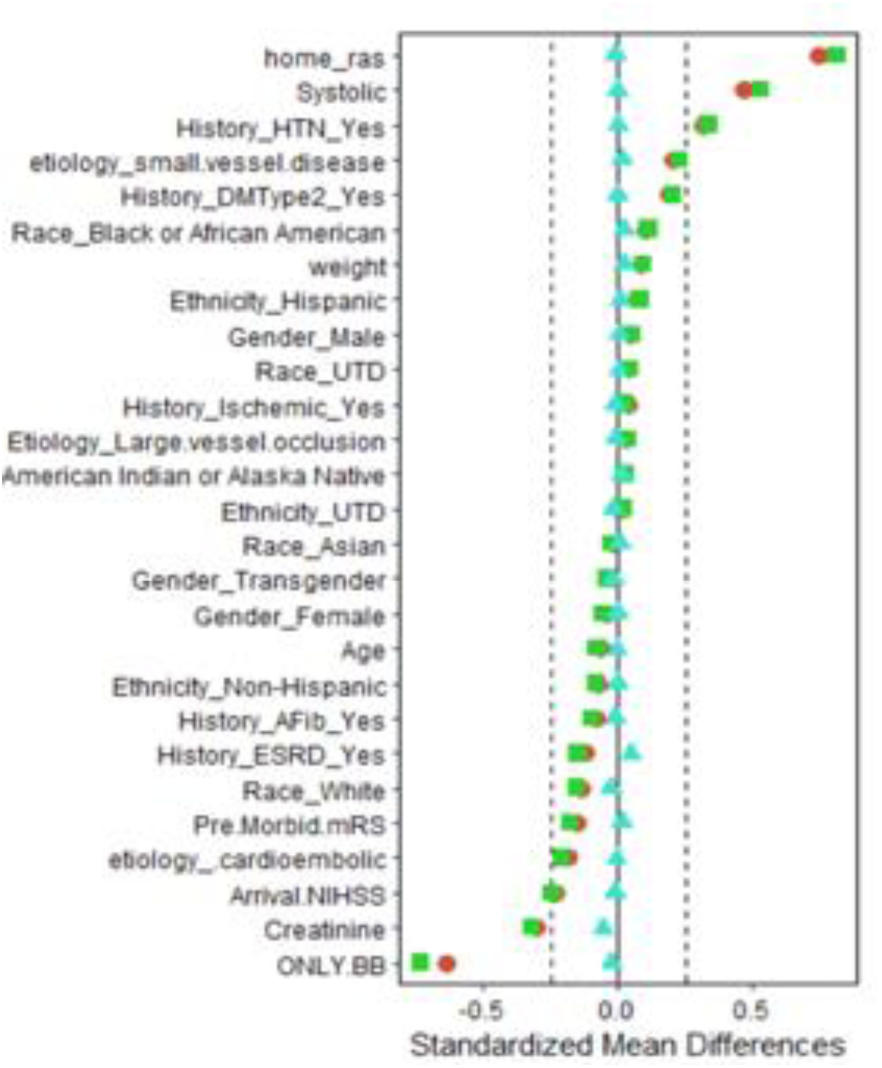
Balance plot for cross sectional analysis

In Figure 3b, we plan to display representative balance plots for longitudinal IPTW and IPCW. It is noteworthy that while balance was effectively attained for the majority of variables, there were residual imbalances observed in patients using RAS as their home medications (home RAS), previous day RAS, and same day creatinine levels. As a result, we included these three variables as covariates in the final model.

**Figure 3b.**
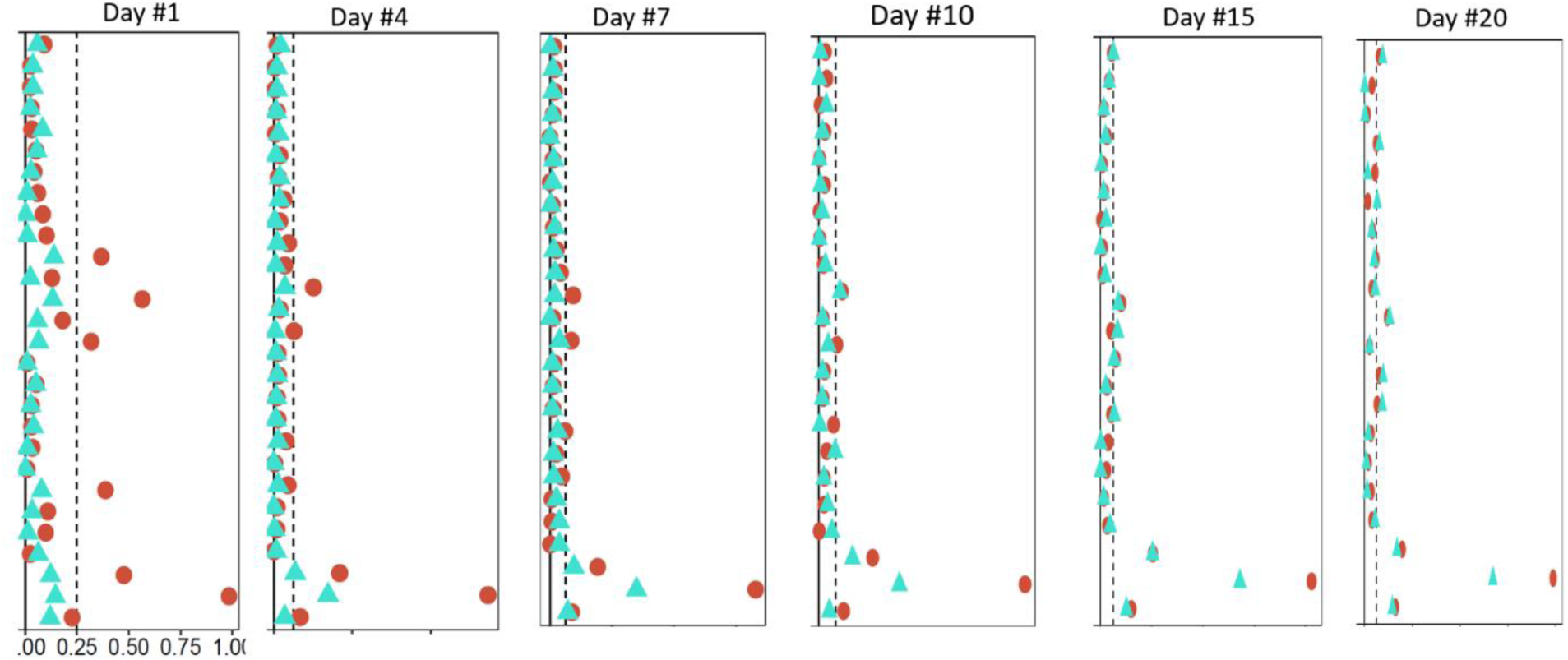
Balance plot for longitudinal analysis using RAS home medication, previous day RAS and daily creatinine levels.

### Cross Sectional Analysis

The results from the IPTW logistic regression model are shown in Table 2. The coefficient for RAS intake indicated a noticeable yet statistically non-significant association with the odds of mortality (OR 0.13, 95% CI 0.01-1.36, p=0.123).

**Table 2.** Cross-Sectional Regression Analysis Results Summary.

| Association | OR | 95% CI | B estimate (coefficient) | SE | p-value |
| --- | --- | --- | --- | --- | --- |
| RAS inhibitors: | 0.13 | 0.01-1.36 | -2.040 | 0.61 | 0.123 |
| Beta Blocker: | 1.07 | 0.73-1.56 | 0.067 | 0.28 | 0.546 |
| Baseline Creatinine: | 1.00 | 0.99-1.02 | 0.000 | 0.01 | 0.901 |
| ESRD: | 2.07 | 0.95-4.53 | 0.729 | 0.48 | 0.067 |
| Interaction |  |  |  |  |  |
| RAS*Beta Blocker: | 0.19 | 0.02-1.52 | -1.659 | 0.47 | 0.314 |
| RAS*Creatinine | 0.78 | 0.46-1.35 | -0.248 | 0.26 | 0.135 |
| RAS*ESRD: | 0.80 | 0.06-10.83 | -0.223 | 0.34 | 0.819 |

### Cross Sectional Analysis -G- Computation

To calculate the unconditional ATE of RAS intake, we employed the G-computation method. Overall, RAS inhibitors significantly reduce risk compared to the ‘No RAS’ group (Risk 0.0281 vs. 0.0913, RD 0.0631 or 6.31%, p < 0.001). Subgroup analysis reveals distinct patterns, with individuals having creatinine levels below and above 1.3 exhibiting statistically significant RD (RD −0.0510 vs. −0.0895, p-value <0.001), and a significant difference in paired comparison (−0.0385 or 3.85%, p-value <0.001). (Table 3).

**Table 3.**
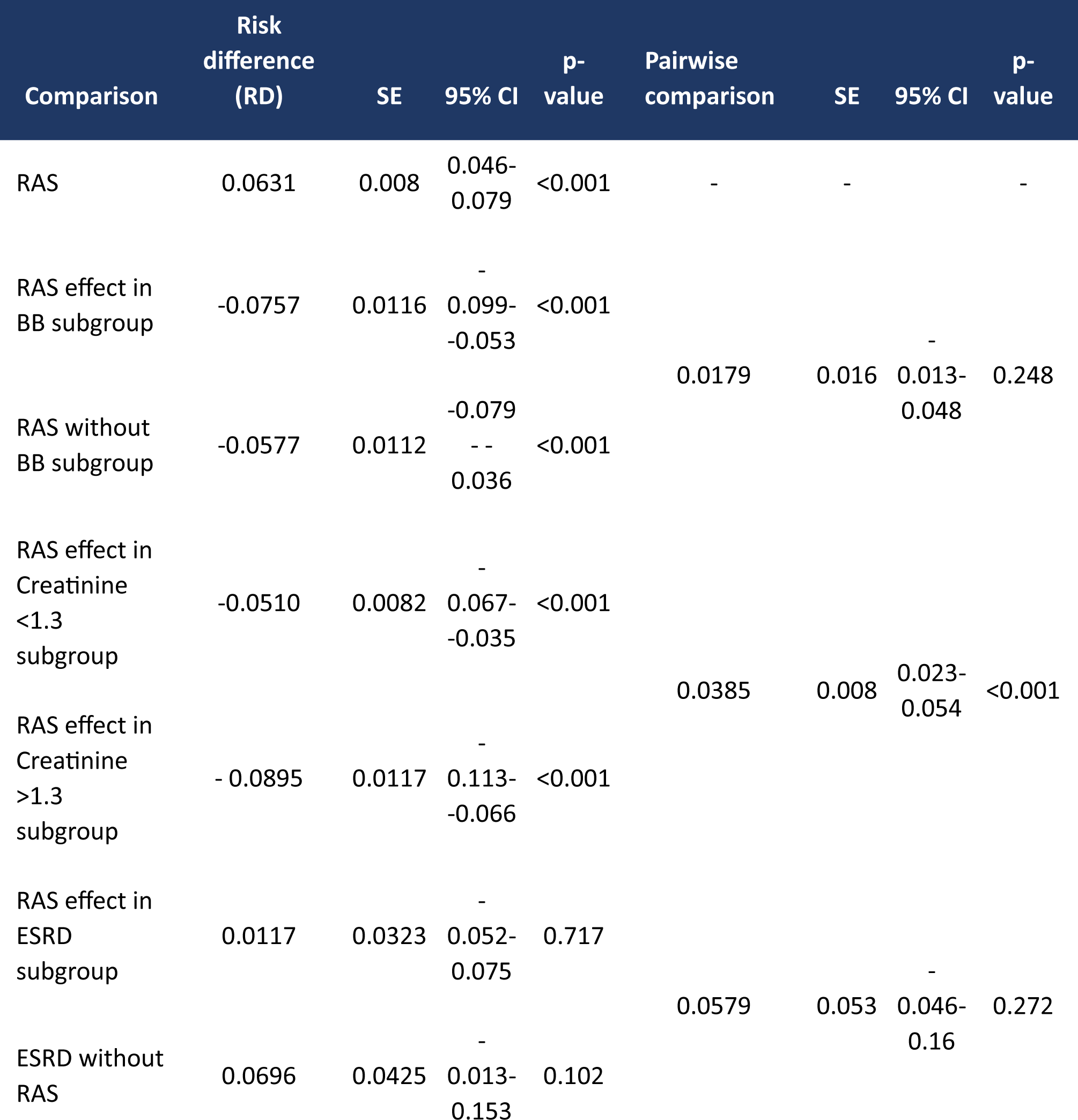
G-Computation Table: Risk Differences and Subgroup Analysis of RAS Inhibitors on Mortality. Cross sectional analysis. Serum creatinine was dichotomized arbitrarily at the 1.3 threshold for the purpose of evaluating the impact of RAS on increasing serum creatinine levels.

### Longitudinal Analysis

In this section, we present a comprehensive analysis of the impact of RAS medication administration on patient outcomes over a 20-day period. To provide a clear overview of our findings, we first present Table 4 below, which summarizes key information, including the number of patients receiving RAS medication, mortality rates for each medication group, and mean creatinine levels as an indicator of kidney function.

**Table 4.**
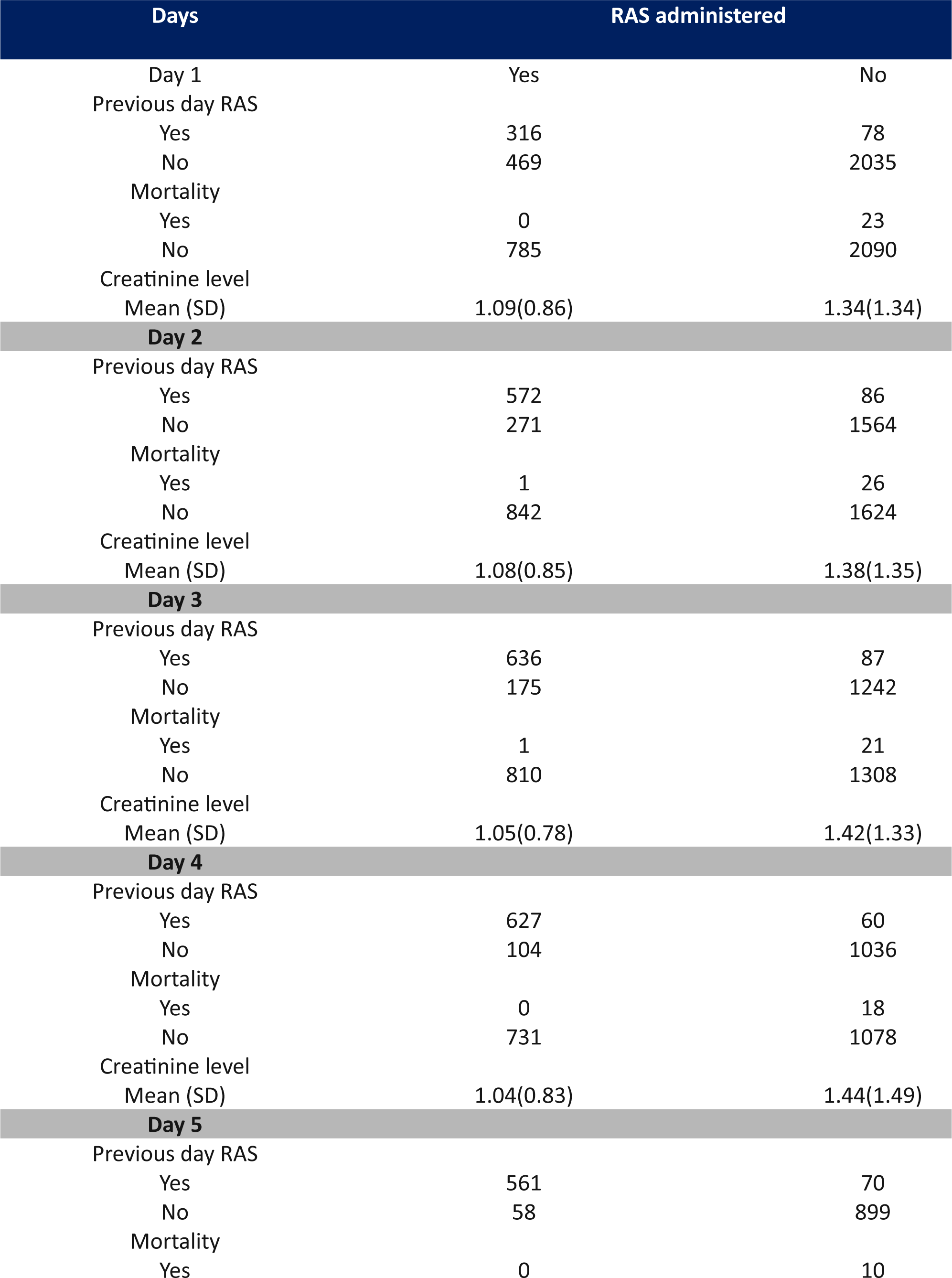
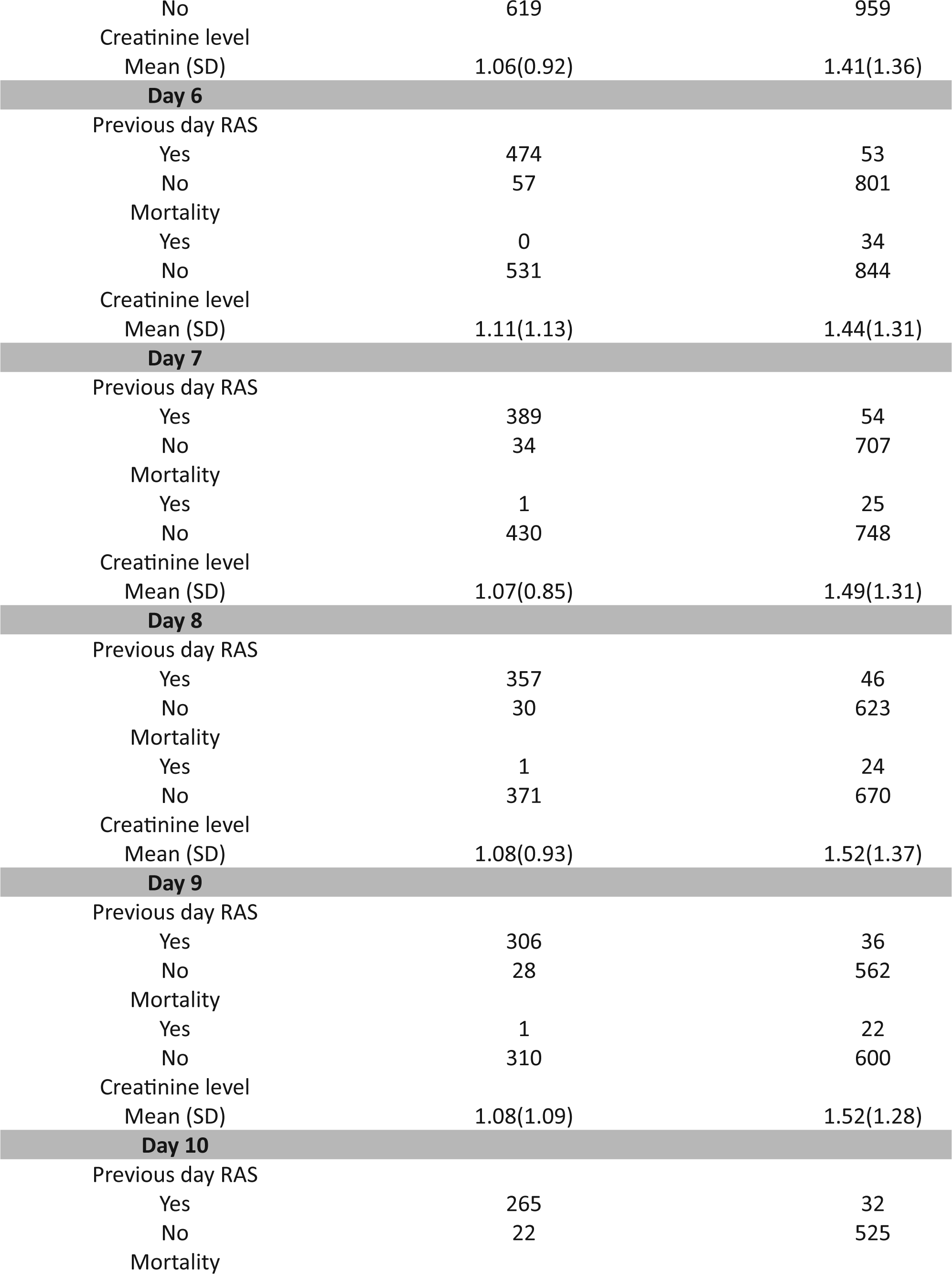
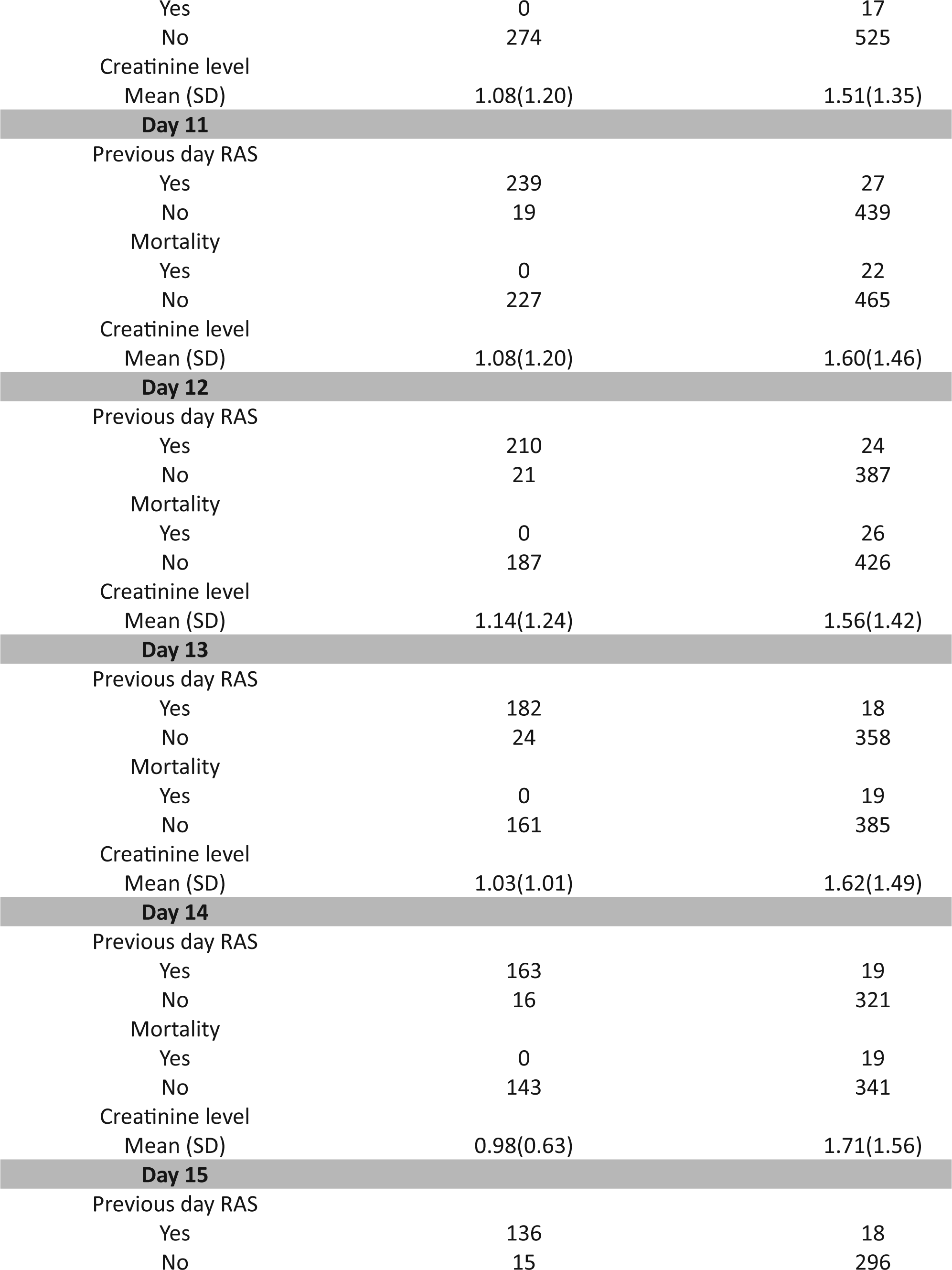
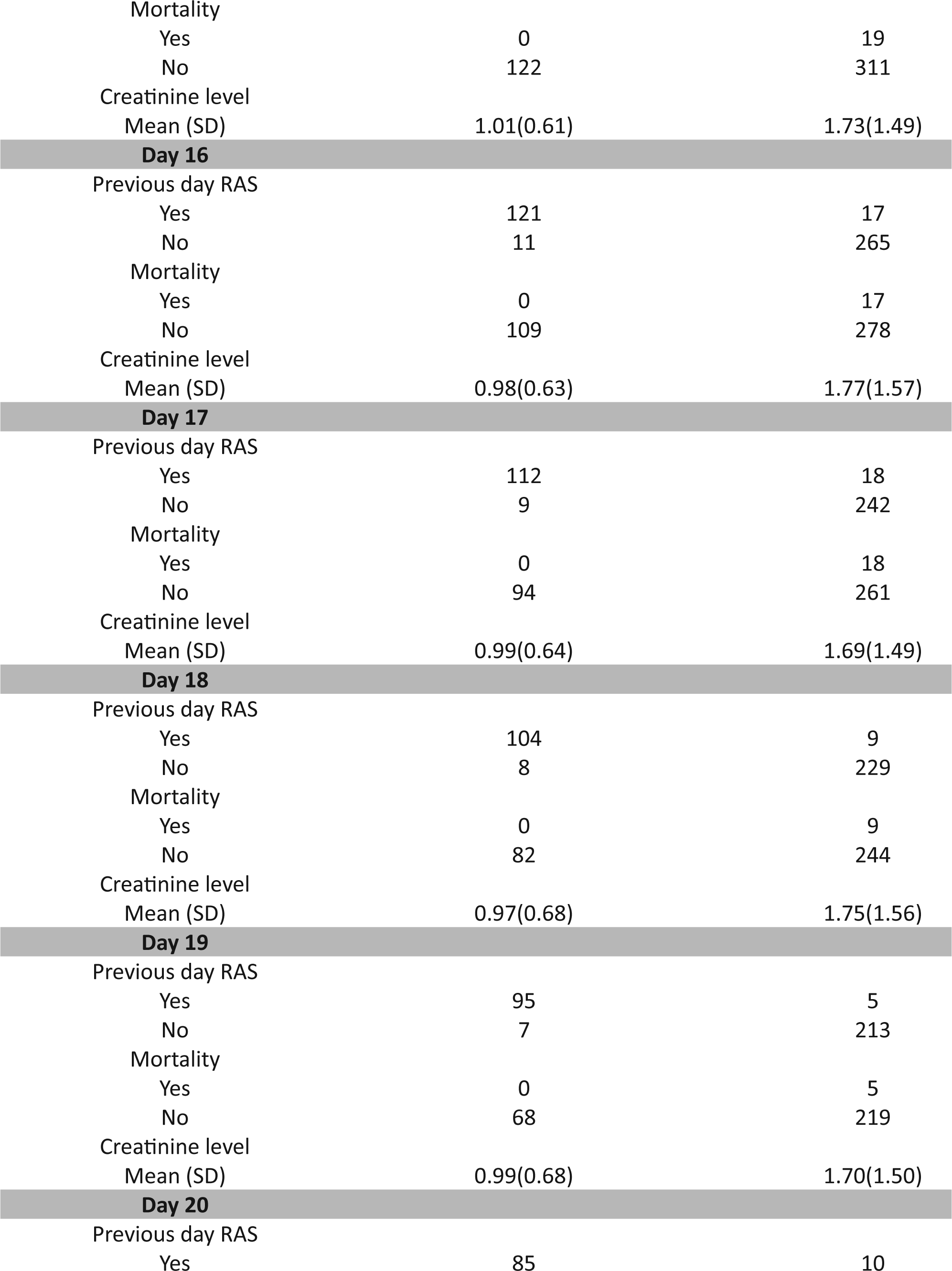
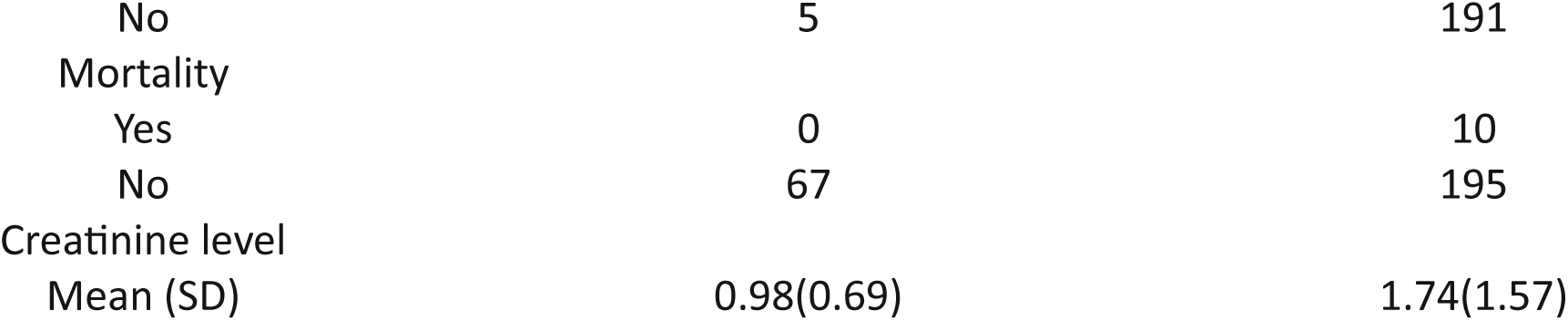
Summary of RAS medication and Clinical Outcomes Over 20 days.

| Days |  | RAS administered |  |
| --- | --- | --- | --- |
| Day 1 |  | Yes | No |
| Previous day RAS |  |  |  |
| Yes |  | 316 | 78 |
| No |  | 469 | 2035 |
| Mortality |  |  |  |
| Yes |  | 0 | 23 |
| No |  | 785 | 2090 |
| Creatinine level |  |  |  |
| Mean (SD) |  | 1.09(0.86) | 1.34(1.34) |
| Day 2 |  |  |  |
| Previous day RAS |  |  |  |
| Yes |  | 572 | 86 |
| No |  | 271 | 1564 |
| Mortality |  |  |  |
| Yes |  | 1 | 26 |
| No |  | 842 | 1624 |
| Creatinine level |  |  |  |
| Mean (SD) |  | 1.08(0.85) | 1.38(1.35) |
| Day 3 |  |  |  |
| Previous day RAS |  |  |  |
| Yes |  | 636 | 87 |
| No |  | 175 | 1242 |
| Mortality |  |  |  |
| Yes |  | 1 | 21 |
| No |  | 810 | 1308 |
| Creatinine level |  |  |  |
| Mean (SD) |  | 1.05(0.78) | 1.42(1.33) |
| Day 4 |  |  |  |
| Previous day RAS |  |  |  |
| Yes |  | 627 | 60 |
| No |  | 104 | 1036 |
| Mortality |  |  |  |
| Yes |  | 0 | 18 |
| No |  | 731 | 1078 |
| Creatinine level |  |  |  |
| Mean (SD) |  | 1.04(0.83) | 1.44(1.49) |
| Day 5 |  |  |  |
| Previous day RAS |  |  |  |
| Yes |  | 561 | 70 |
| No |  | 58 | 899 |
| Mortality |  |  |  |
| Yes |  | 0 | 10 |

**Table 4.** Summary of RAS medication and Clinical Outcomes Over 20 days.
|  |  |  |
| --- | --- | --- |
| No | 619 | 959 |
| Creatinine level |  |  |
| Mean (SD) | 1.06(0.92) | 1.41(1.36) |
| **Day 6** |  |  |
| Previous day RAS |  |  |
| Yes | 474 | 53 |
| No | 57 | 801 |
| Mortality |  |  |
| Yes | 0 | 34 |
| No | 531 | 844 |
| Creatinine level |  |  |
| Mean (SD) | 1.11(1.13) | 1.44(1.31) |
| **Day 7** |  |  |
| Previous day RAS |  |  |
| Yes | 389 | 54 |
| No | 34 | 707 |
| Mortality |  |  |
| Yes | 1 | 25 |
| No | 430 | 748 |
| Creatinine level |  |  |
| Mean (SD) | 1.07(0.85) | 1.49(1.31) |
| **Day 8** |  |  |
| Previous day RAS |  |  |
| Yes | 357 | 46 |
| No | 30 | 623 |
| Mortality |  |  |
| Yes | 1 | 24 |
| No | 371 | 670 |
| Creatinine level |  |  |
| Mean (SD) | 1.08(0.93) | 1.52(1.37) |
| **Day 9** |  |  |
| Previous day RAS |  |  |
| Yes | 306 | 36 |
| No | 28 | 562 |
| Mortality |  |  |
| Yes | 1 | 22 |
| No | 310 | 600 |
| Creatinine level |  |  |
| Mean (SD) | 1.08(1.09) | 1.52(1.28) |
| **Day 10** |  |  |
| Previous day RAS |  |  |
| Yes | 265 | 32 |
| No | 22 | 525 |
| Mortality |  |  |
| Yes | 0 | 17 |
| No | 274 | 525 |
| Creatinine level |  |  |
| Mean (SD) | 1.08(1.20) | 1.51(1.35) |
| **Day 11** |  |  |
| Previous day RAS |  |  |
| Yes | 239 | 27 |
| No | 19 | 439 |
| Mortality |  |  |
| Yes | 0 | 22 |
| No | 227 | 465 |
| Creatinine level |  |  |
| Mean (SD) | 1.08(1.20) | 1.60(1.46) |
| **Day 12** |  |  |
| Previous day RAS |  |  |
| Yes | 210 | 24 |
| No | 21 | 387 |
| Mortality |  |  |
| Yes | 0 | 26 |
| No | 187 | 426 |
| Creatinine level |  |  |
| Mean (SD) | 1.14(1.24) | 1.56(1.42) |
| **Day 13** |  |  |
| Previous day RAS |  |  |
| Yes | 182 | 18 |
| No | 24 | 358 |
| Mortality |  |  |
| Yes | 0 | 19 |
| No | 161 | 385 |
| Creatinine level |  |  |
| Mean (SD) | 1.03(1.01) | 1.62(1.49) |
| **Day 14** |  |  |
| Previous day RAS |  |  |
| Yes | 163 | 19 |
| No | 16 | 321 |
| Mortality |  |  |
| Yes | 0 | 19 |
| No | 143 | 341 |
| Creatinine level |  |  |
| Mean (SD) | 0.98(0.63) | 1.71(1.56) |
| **Day 15** |  |  |
| Previous day RAS |  |  |
| Yes | 136 | 18 |
| No | 15 | 296 |
| Mortality |  |  |
| Yes | 0 | 19 |
| No | 122 | 311 |
| Creatinine level |  |  |
| Mean (SD) | 1.01(0.61) | 1.73(1.49) |
| **Day 16** |  |  |
| Previous day RAS |  |  |
| Yes | 121 | 17 |
| No | 11 | 265 |
| Mortality |  |  |
| Yes | 0 | 17 |
| No | 109 | 278 |
| Creatinine level |  |  |
| Mean (SD) | 0.98(0.63) | 1.77(1.57) |
| **Day 17** |  |  |
| Previous day RAS |  |  |
| Yes | 112 | 18 |
| No | 9 | 242 |
| Mortality |  |  |
| Yes | 0 | 18 |
| No | 94 | 261 |
| Creatinine level |  |  |
| Mean (SD) | 0.99(0.64) | 1.69(1.49) |
| **Day 18** |  |  |
| Previous day RAS |  |  |
| Yes | 104 | 9 |
| No | 8 | 229 |
| Mortality |  |  |
| Yes | 0 | 9 |
| No | 82 | 244 |
| Creatinine level |  |  |
| Mean (SD) | 0.97(0.68) | 1.75(1.56) |
| **Day 19** |  |  |
| Previous day RAS |  |  |
| Yes | 95 | 5 |
| No | 7 | 213 |
| Mortality |  |  |
| Yes | 0 | 5 |
| No | 68 | 219 |
| Creatinine level |  |  |
| Mean (SD) | 0.99(0.68) | 1.70(1.50) |
| **Day 20** |  |  |
| Previous day RAS |  |  |
| Yes | 85 | 10 |
| No | 5 | 191 |
| Mortality |  |  |
| Yes | 0 | 10 |
| No | 67 | 195 |
| Creatinine level |  |  |
| Mean (SD) | 0.98(0.69) | 1.74(1.57) |

Following this detailed longitudinal analysis, we transition to the results from the IPW multiperiod logit model in the subsequent section. Within this model, we identify a highly significant coefficient of −3.71 for the daily RAS variable (p < 0.001), signifying a robust association with the outcome. (Table 5) For the computation of the ATE, G-computation was employed.

**Table 5.**
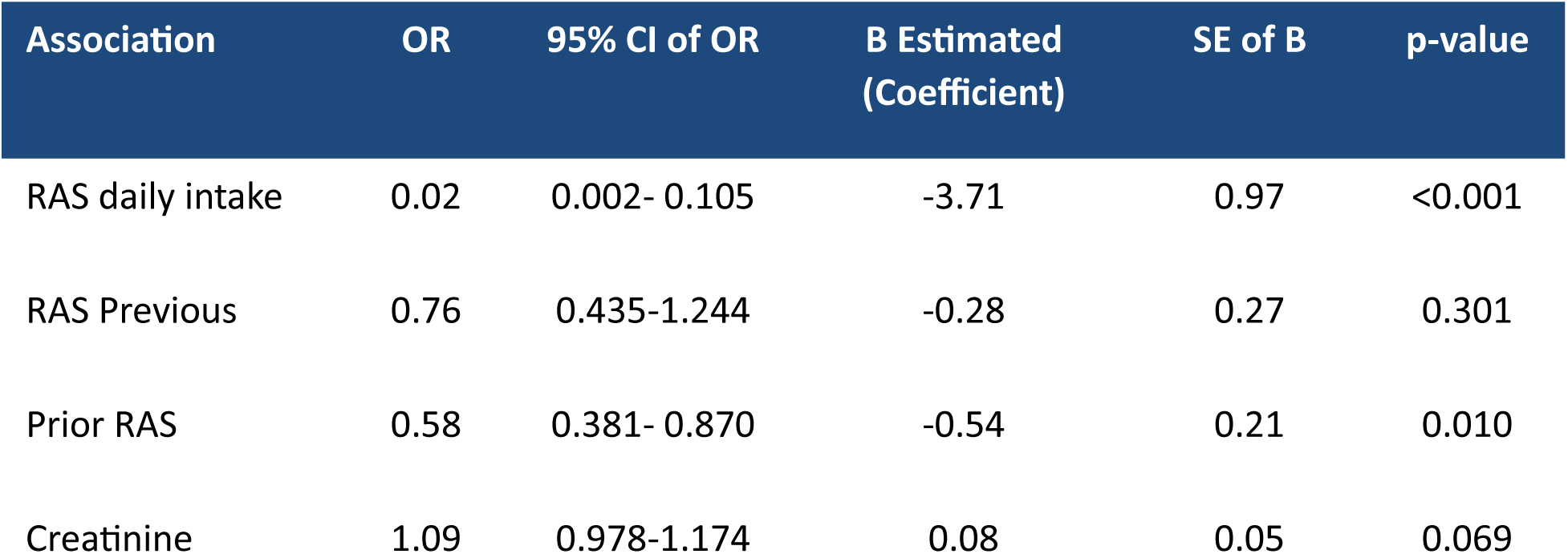
Discrete time survival model: Logistic Regression.

| Association | OR | 95% CI of OR | B Estimated<br>(Coefficient) | SE of B | p-value |
| --- | --- | --- | --- | --- | --- |
| RAS daily intake | 0.02 | 0.002- 0.105 | -3.71 | 0.97 | <0.001 |
| RAS Previous | 0.76 | 0.435-1.244 | -0.28 | 0.27 | 0.301 |
| Prior RAS | 0.58 | 0.381- 0.870 | -0.54 | 0.21 | 0.010 |
| Creatinine | 1.09 | 0.978-1.174 | 0.08 | 0.05 | 0.069 |

### Longitudinal Analysis G-Computation

In our G-Computation analysis, we observed significant effect of daily RAS on mortality. Patients who received RAS intake exhibited a daily risk (i.e., hazard) of 0.000237, whereas patients with no RAS intake had a daily risk of 0.00955. The risk difference (RD) was estimated to be 0.00931, equivalent to a 0.93% daily reduction in mortality, with SE=0.00106, 95% CI [0.00722, 0.0114], a p-value of <0.001. These findings highlight substantial differences between the groups, particularly in terms of the hazard rate and the daily mortality reduction.

### Absolute Risk Reduction (ARR) and Number Needed to Treat (NNT)

In the preliminary data analysis, a substantial absolute risk reduction (ARR) of 10% was observed, corresponding to a number needed to treat (NNT) of 10. The cross-sectional analysis revealed a moderate ARR of 6.3%, with an NNT of 16. Conversely, the longitudinal analysis unveiled a smaller daily ARR of 0.9%, highlighting the cumulative effect over time.

## Discussion

In our comprehensive study, we investigated the impact of RAS inhibitors (ACE inhibitors or ARBs) compared to non-RAS antihypertensives on mortality in AIS patients. Notably, a significant proportion of patients had been on RAS medications at home, and all were resumed with their home medications upon admission to the hospital. This presented a unique challenge and potential confounder, which we diligently addressed through advanced statistical techniques. Our initial analysis, which revealed a short median duration of RAS inhibitor use (4 days), underscored the importance of understanding the resumption of RAS home medications.^36^ To enhance the robustness of our findings and address potential biases stemming from this resumption, we employed advanced statistical methods, including imputation techniques, IPTW, regression analysis, and G-computation. These approaches allowed us to thoroughly adjust for the complexities introduced by resuming home medications and strengthened our ability to draw reliable conclusions. In our cross-sectional analysis, we observed a significant reduction in mortality associated with RAS inhibitors, with no synergistic effect when combined with BBs. Subgroup analysis suggested potential benefits, particularly in patients with elevated creatinine levels and possibly ESRD. Transitioning to longitudinal analysis allowed us to track RAS intervention and mortality outcomes over time, providing more reliable estimates. We quantified the impact using absolute risk reduction (ARR) and number needed to treat (NNT), highlighting cumulative effects over time. Beyond blood pressure management, our study emphasized the broader benefits of RAS inhibitors on renal outcomes.^7,16,17^ While comparing our findings with the SCAST^37^ trial revealed differences in patient selection and outcomes, our study’s specificity to AIS patients and in-hospital mortality emphasizes its unique contributions. Limitations of our study include its retrospective design, potential confounding, single-center setting, and reliance on electronic medical records. Future prospective studies are essential to confirm our findings. In conclusion, our study suggests a potential daily reduction in mortality (0.9%) associated with RAS inhibitors in AIS patients. To comprehensively assess their impact, particularly on functional outcomes, rigorous longitudinal analysis is vital. This investigation will provide a more comprehensive evaluation of RAS inhibitors’ potential to enhance outcomes in acute ischemic stroke patients.

## Data Availability

We acknowledge and confirm that all data and materials referred to in this manuscript are available upon request.

